# Percutaneous coronary intervention has better in-hospital outcomes in severe renal failure patients

**DOI:** 10.1101/2023.05.03.23289446

**Authors:** Renxi Li, Qianyun Luo

## Abstract

**Background:** In patients with severe renal failure, coronary artery bypass surgery (CABG) was found to have better long-term outcomes than percutaneous coronary intervention (PCI). This study aims to provide a retrospective analysis of short-term in-hospital peri-operative outcomes between PCI and CABG in patients with severe renal failure.

**Methods:** Patients who underwent CABG and PCI in Q4 2015-2020 were identified in National Inpatient Sample (NIS) database. Patients of age<40 were excluded for congenital heart defects. Patients with comorbidity of severe renal failure were included. Between patients undergoing PCI and CABG, preoperative variables were compared and corrected in multivariable logistic regression examining their in-hospital peri-operative outcomes. Adjusted odds ratios (aOR) were estimated for mortality and morbidities.

**Results:** In NIS, 4,512 CABG and 13,242 PCI cases were identified. Compared to CABG, patients who underwent PCI had lower mortality (4.24% vs 5.47%, aOR 0.725, p<0.0001) and lower morbidity including heart failure (0.14% vs 2.97%, aOR 0.044, p<0.0001), stroke (0.12% vs 0.47%, aOR 0.264, p<0.0001), respiratory complications (0.51% vs 7.34%, aOR 0.063, p<0.0001), renal complications (0.05% vs 0.49%, aOR 0.075, p<0.0001), acute kidney injury (8.67% vs 12.66%, aOR 0.646, p<0.0001), deep wound complications (0.02% vs 0.35%, aOR 0.042, p < 0.0001), shock (0.22% vs 0.95%, aOR 0.228, p<0.0001), and length of in-hospital stay over 7 days (24.75% vs 73.52%, aOR 0.106, p<0.0001).

**Conclusion:** NIS is a comprehensive database of nationwide providers, providing robust power in analysis. In patients with severe renal failure, PCI offers an advantage over CABG in terms of short-term in-hospital perioperative outcomes.

## Introduction

Coronary Artery Disease (CAD) is a leading cause of death in the US. When indicated for coronary revascularization, coronary artery bypass grafting (CAGB) and percutaneous coronary intervention (PCI) are the two most modalities ^1^. Severe renal failure, which includes stage 5 chronic kidney disease (CKD) and end-stage renal disease (ESRD), is a risk factor for developing CAD ^2^. There are targeted randomized controlled trials such as EXCEL (Evaluation of XIENCE Versus Coronary Artery Bypass Surgery for Effectiveness of Left Main Revascularization) ^3^. However, a comprehensive randomized controlled trial comparing the outcomes of CABG and PCI in patients with severe renal failure is still lacking and there is still ongoing debate on which modality is better ^4^.

Previous meta-analysis found CABG offers better long-term outcomes than PCI in terms of all-cause and cardiac mortality while short-term mortality is higher in CABG ^5^. However, there is a lack of research focused on other short-term in-hospital outcomes, such as infection, acute kidney injury, venous thromboembolism/pulmonary embolism. it is crucial to emphasize the significance of these short-term in-hospital outcomes in clinical decision-making as they can help physicians provide prompt in-patient care to avoid further complications.

This study aims to provide a comprehensive analysis of the in-hospital peri-operative outcomes between PCI and CABG in patients with severe renal failure. This study will be retrospective based on National/Nationwide Inpatient Sample (NIS), which is the largest inpatient database that records 20% of all discharges from hospitals in the US. As many previous studies were based on local institutional data, this study will provide considerably more statistical power to assess the peri-operative outcomes.

## Patients and Methods

Patients who underwent CABG and PCI in the last quarter of 2015-2020 were identified in the NIS database by ICD10-PCS (CABG: 0210xxx; PCI: 02703xx, 02713xx, 02723xx, and 02733xx, excluding 027x3Tx and 027x3Zx). Patients of age less than 40 were excluded for congenital heart defects. Comorbidities were defined by Elixhauser measure ^6^. Patients with severe renal failure comorbidity (Stage 5 CKD and ESRD) were included.

Fisher’s exact test was used to test for categorical variables while a two-sample independent t-test was used for continuous variables. Preoperative differences between patients undergoing PCI and CABG were noted. Perioperative outcomes were examined by univariate analysis and then adjusted for preoperative differences using multivariable logistic regression, where preoperative variables with noted differences (p-value < 0.1) were included in the regression. Adjusted odds ratios (aOR) were estimated between PCI and CABG groups for mortality, heart failure, stroke, myocardial infarction (MI), major adverse cardiovascular events (MACE), respiratory complications, pulmonary embolism (PE), venous thromboembolism (VTE), renal complications, acute kidney injury, bleeding, superficial/deep wound, sepsis, shock, length of stay>7 days, and transfer.

All statistical analyses were performed using SAS (version 9.4). Given the usage of retrospective, de-identified NIS data, this study was exempted from IRB approval.

## Results

There were 4,512 CABG and 13,242 PCI cases with severe renal failure identified. The comorbidities and demographics of the patients are summarized in Table 1. Severe renal failure patients who underwent PCI were more likely to be Black, female, older age, and with comorbidities of lymphoma, malignant solid tumor without metastasis, dementia, diabetes, severe liver disease, chronic lung disease, hypothyroidism, and valve disease. In contrast, patients who underwent CABG are more likely to be Hispanic, Asian, male, and have heart failure and complicated hypertension.

**Table 1.** Comorbidities and demographics of patients with severe renal failure who underwent CABG or PCI between the last quarter of 2015 and 2020 in NIS database.

|  | **CABG (n=4,512)** | **PCI (n=13,242)** | **p-value** |
| --- | --- | --- | --- |
| **Race, n (%)** |  |  |  |
| White | 2109 (46.74%) | 6180 (46.67%) | 0.9312 |
| Black | 900 (19.95%) | 3158 (23.85%) | **<.0001** |
| Hispanic | 815 (18.06%) | 2159 (16.3%) | **0.0069** |
| Asian | 330 (7.31%) | 789 (5.96%) | **0.0014** |
| Native Americans | 55 (1.22%) | 175 (1.32%) | 0.6477 |
| Others | 174 (3.86%) | 474 (3.58%) | 0.4080 |
| **Sex, n (%)** |  |  |  |
| Male sex | 3184 (70.57%) | 8063 (60.88%) | **<.0001** |
| Female sex | 1328 (29.43%) | 5179 (39.11%) | **<.0001** |
| **Age, mean ± SD, years old** | **62.42 ± 10.32** | **64.87 ± 11.43** | **<.0001** |
| **Comorbidities, n (%)** |  |  |  |
| AIDS | 42 (0.93%) | 128 (0.97%) | 0.9295 |
| Alcohol use | 42 (0.93%) | 111 (0.84%) | 0.5759 |
| Autoimmune Disease | 130 (2.88%) | 374 (2.82%) | 0.8356 |
| Lymphoma | 16 (0.35%) | 92 (0.69%) | **0.0104** |
| Leukemia | 18 (0.4%) | 42 (0.32%) | 0.4572 |
| Metastatic cancer | 9 (0.2%) | 51 (0.39%) | 0.0738 |
| Solid tumor without metastasis, in situ | 2 (0.04%) | 3 (0.02%) | 0.6067 |
| Solid tumor without metastasis, malignant | 44 (0.98%) | 181 (1.37%) | 0.0449 |
| Cerebral vascular disease | 161 (3.57%) | 518 (3.91%) | 0.3227 |
| Heart Failure | 7 (0.16%) | 1 (0.01%) | **0.0004** |
| Dementia | 46 (1.02%) | 345 (2.61%) | **<.0001** |
| Depression | 410 (9.09%) | 1324 (10%) | 0.0766 |
| Diabetes without chronic complications | 246 (5.45%) | 829 (6.26%) | 0.051 |
| Diabetes with chronic complications | 3226 (71.5%) | 9175 (69.28%) | **0.0051** |
| Drug abuse | 62 (1.37%) | 166 (1.25%) | 0.5404 |
| Hypertension complicated | 4176 (92.55%) | 11787 (89.01%) | **<.0001** |
| Hypertension uncomplicated | 132 (2.93%) | 419 (3.16%) | 0.4559 |
| Severe liver disease | 18 (0.4%) | 103 (0.78%) | **0.0063** |
| Chronic lung disease | 761 (16.87%) | 2728 (20.6%) | **<.0001** |
| Obesity | 1173 (26%) | 2735 (20.65%) | **<.0001** |
| Paralysis | 131 (2.9%) | 425 (3.21%) | 0.3225 |
| Peripheral vascular disease | 777 (17.22%) | 2202 (16.63%) | 0.3563 |
| Hypothyroidism | 562 (12.46%) | 1850 (13.97%) | **0.0103** |
| Other thyroid disorders | 35 (0.78%) | 104 (0.79%) | 1 |
| Valve disease | 42 (0.93%) | 338 (2.55%) | **<.0001** |

The univariate and multivariable logistic regression comparing the in-hospital perioperative outcomes of CABG and PCI in severe renal failure patients were summarized in Table 2.

**Table 2.** The in-hospital post-CABG and PCI surgical outcomes of patients with severe renal failure from 2015 to 2020 in the NIS database.

|  | **PCI (n = 13243)**<br>No. of case (%) | **CABG (n = 4512)**<br>No. of case (%) | **aOR for**<br>**PCI/CABG, 95%**<br>**CI** | **p-value** |
| --- | --- | --- | --- | --- |
| Mortality | 562 (4.24%) | 247 (5.47%) | 0.725, 0.620-0.847 | **<.0001** |
| Heart failure | 19 (0.14%) | 134 (2.97%) | 0.044, 0.027-0.071 | **<.0001** |
| Stroke | 16 (0.12%) | 21 (0.47%) | 0.264, 0.138-0.507 | **<.0001** |
| Myocardial infarction | 358 (2.7%) | 115 (2.55%) | 1.053, 0.850-1.306 | 0.6348 |
| Respiratory complication | 67 (0.51%) | 331 (7.34%) | 0.063, 0.049-0.083 | **<.0001** |
| Pulmonary embolism | 5 (0.04%) | 1 (0.02%) | 1.704, 0.199-14.587 | 0.6267 |
| Venous thromboembolism | 54 (0.41%) | 24 (0.53%) | 0.758, 0.468-1.229 | 0.2616 |
| Renal failure | 6 (0.05%) | 22 (0.49%) | 0.075, 0.030-0.189 | **<.0001** |
| Acute kidney injury | 1148 (8.67%) | 571 (12.66%) | 0.646, 0.579-0.720 | **<.0001** |
| Bleeding events | 27 (0.2%) | 12 (0.27%) | 0.772, 0.391-1.526 | 0.457 |
| Superficial wound | 142 (1.07%) | 44 (0.98%) | 1.012, 0.718-1.426 | 0.9445 |
| Deep wound | 2 (0.02%) | 16 (0.35%) | 0.042, 0.010-0.182 | **<.0001** |
| Sepsis | 5 (0.04%) | 4 (0.09%) | 0.412, 0.110-1.537 | 0.1866 |
| Shock | 29 (0.22%) | 43 (0.95%) | 0.228, 0.142-0.365 | **<.0001** |
| Length of stay>7 days | 3277 (24.75%) | 3317 (73.52%) | 0.106, 0.098-0.115 | **<.0001** |
| Transfer | 2065 (15.59%) | 1723 (38.19%) | 0.270, 0.250-0.292 | **<.0001** |

Compared to CABG, severe renal failure patients who received PCI had lower mortality (4.24% vs 5.47%, aOR 0.725, p < 0.0001) and had lower morbidity including heart failure (0.14% vs 2.97%, aOR 0.044, p < 0.0001), stroke (0.12% vs 0.47%, aOR 0.264, p < 0.0001), respiratory complications (0.51% vs 7.34%, aOR 0.063, p < 0.0001), renal complications (0.05% vs 0.49%, aOR 0.075, p < 0.0001), acute kidney injury (8.67% vs 12.66%, aOR 0.646, p < 0.0001), deep wound complications (0.02% vs 0.35%, aOR 0.042, p < 0.0001), shock (0.22% vs 0.95%, aOR 0.228, p < 0.0001), and length of in-hospital stay over 7 days (24.75% vs 73.52%, aOR 0.106, p < 0.0001). There was no difference in MI, PE, VTE, bleeding, superficial wound infection, or sepsis.

## Comment

NIS is a comprehensive database of nationwide providers, providing robust power in the analysis in in-hospital outcomes. This study aimed to provide a comprehensive comparison of PCI and CABG in terms of their in-hospital peri-operative outcome in patients with severe renal failure.

CABG is more invasive and had worse immediate outcomes, as shown in the study. Thus, patients in more unfavorable conditions such as being at an older age, having diabetes, malignancy, liver and lung diseases tended to undergo PCI. However, since CABG offers a better long-term prognosis, it is a better option for patients with a history of heart failure and complicated hypertension ^7^.

PCI has less morality, heart failure, and stroke, which are consistent with the previous meta-analysis ^5^. Pulmonary complications are unavoidable in CABG, which is confirmed by the presence of more immediate respiratory complications in CABG patients in this study ^8^. A randomized control trial revealed less AKI/acute-on-chronic kidney disease in severe renal failure patients who underwent PCI for left main coronary artery revascularization ^3^. This study further shows in all severe renal failure patients who underwent PCI, there is less risk for acute-on-chronic kidney diseases. The rate of VTE and PE were low in both CABG and PCI and there is no difference in their occurrence, which maybe account for by both adequate anti-coagulant management in both modalities. CABG, as a more complex and invasive procedure, not surprisingly required longer stay in the hospital, higher transfer rate to other healthcare facilities, and has more perioperative deep wound infection and shook.

There are various limitations of this study. Previous studies demonstrated intraoperative factors, including affected stenosis diameter, coronary segment, the dominance of the coronary arteries, presence of lesions, and diffusion to small vessels, that are relevant to successful revascularization and may affect morbidity and mortality but are not recorded in NIS ^1,9^. Also, numerous factors cannot be controlled for retrospective studies, such as the accuracy of the record and selection bias of the participating hospitals in NIS. Thus, comprehensive randomized studies are needed in the future the assess these coronary revascularization modalities in patients with severe renal failure.

Overall, while the literature shows CABG offers better long-term outcomes in CAD patients with severe renal failure, this study demonstrated that short-term outcomes are superior in PCI not only in mortality and cardiac complications but also in terms of pulmonary, renal, and wound complications. This study is thus instructive for providers to consider the potential risks and benefits of each intervention for patients with severe renal failure. For example, high-risk renal failure patients might prefer the early benefit of PCI. In addition, this study provides insight on perioperative management, where in patients with severe renal failure, besides cardiac complications, acute-on-chronic kidney disease and pulmonary complications should be continuously monitored, especially in patients with relevant comorbidities.

## Data Availability

All data produced in the present study are available upon reasonable request to the authors.

## Acknowledgments

The authors acknowledge the guidance and support of colleagues and reviewers who provided useful feedback throughout the work.

